# The bioavailability of compounded and generic rapamycin in normative aging individuals: A retrospective study and review with clinical implications

**DOI:** 10.1101/2024.08.12.24311432

**Authors:** Girish Harinath, Virginia Lee, Andy Nyquist, Mauricio Moel, Maartje Wouters, Jesper Hagemeier, Brandon Verkennes, Colleen Tacubao, Krister Kauppi, Stefanie L. Morgan, Anar Isman, Sajad Zalzala

## Abstract

Rapamycin, also known as sirolimus, has demonstrated great potential for application in longevity medicine. However, the bioavailability of generic and compounded rapamycin at longevity doses in normative aging individuals remains unknown. We conducted a retrospective, real-world study determining the 24-hour blood rapamycin levels to establish the relative bioavailability, dose-to-blood level linearity and inter-individual heterogeneity in a normative aging cohort. Participants received either compounded rapamycin (n = 23, dosages 5, 10, or 15 mg) or generic rapamycin (n= 44, dosages 2, 3, 6, or 8 mg) once per week, and were asked to obtain a sirolimus level blood draw 24 hours after dose self-administration. Similar blood rapamycin levels and a linear dose-to-blood level relationship were observed for both formulations, although a higher bioavailability per milligram of rapamycin was noted for the generic formulation (compounded averaged 0.287 (28.7%) bioavailability relative to generic rapamycin in (ng/mL) / mg rapamycin). While substantial inter-individual heterogeneity in blood rapamycin levels was observed for both formulations, repeat tests for individuals demonstrated high test-retest reliability. As we detected no significant association between bioavailability and measures of body mass index (BMI), sex, age, or length of time taking rapamycin, we suggest that individualized dosing and routine monitoring of blood rapamycin levels should be applied to ensure optimal longevity efficacy. Finally, we contextualize our data with a brief review of the literature on the currently available knowledge of rapamycin’s bioavailability in normative aging populations, and provide implications for the clinical use of rapamycin in longevity medicine moving forward.

## Introduction

The number of people aged >65 years is expected to reach 2 billion by 2050, which will present significant challenges given the evidence that biological aging is the biggest risk factor for age-related chronic diseases such as cancer, cardiovascular disease, and neurodegenerative diseases [1-3]. The high comorbidity of age-related diseases in elderly individuals limits the benefit that can be obtained by targeting each chronic disease individually. As such, it is essential to implement preventative healthcare strategies that address the biology of aging to limit the rise of multi-morbidity [4, 5].

The translational geroscience field has placed substantial focus on assessing gerotherapeutics targeting the molecular mechanisms underlying biological aging, with the aim of improving healthy longevity [6]. In the past 15 years, several gerotherapeutics have shown efficacy in improving healthy lifespan in preclinical models; however, whether these therapeutics can delay or even prevent multiple age-related diseases in humans remains poorly understood. To address these gaps, translational geroscientists are evaluating US Food and Drug Administration (FDA)-approved interventions for their potential to mitigate age-related decline and be repurposed as gerotherapeutic interventions. Among these, the small molecule rapamycin has been most broadly recognized to hold significant translational promise [7-9].

Rapamycin inhibits the mechanistic target of rapamycin (mTOR), a serine/threonine kinase composed of two functionally distinct complexes, mTOR complex 1 (mTORC1) and 2 (mTORC2). The activities and substrate specificities of mTORC1/2 are regulated by complex co-factors to collectively serve as a molecular control for the maintenance of cellular homeostasis [10]. Potent chemical inhibition of mTORC1 by rapamycin has been extensively characterized, and has demonstrated considerable efficacy in preclinical studies for addressing age-related diseases such as cancer, cardiovascular disease, neurodegenerative disease, and sarcopenia [11-13]. This has spurred significant interest in rapamycin and its derivatives as gerotherapeutics, and early studies in companion animals, non-human primates, and small cohorts of aged individuals have shown promising results [14, 15]. However, further investigation in longitudinal, randomized controlled trials (RCTs) in large and diverse normative aging cohorts is necessary to obtain a more comprehensive understanding of rapamycin’s potential for longevity medicine.

Several clinical trials assessing the effects of various dosing and regimen schedules of rapamycin on age-related decline are currently underway in healthy adults (such as the PEARL trial NCT04488601) [14]. These are of importance to the geroscience field, as existing data on dosing regimens that extend lifespan have only come from model organisms. Thus, our current knowledge cannot be directly translated to human doses, as factors such as interspecies differences in bioavailability, half-life, clearance, plasma protein binding, and tissue distribution play critical roles in functional outcomes [14, 16]. As such, understanding these factors in normative aging individuals will be essential for the effective application of rapamycin as a gerotherapeutic in the future.

The pharmacokinetics (PK) of rapamycin have been thoroughly characterized in relation to the FDA-approved uses for rapamycin, such as anti-rejection for kidney transplants, and treatment of tuberous sclerosis, solid tumors, and lymphomas [17-20]. However, the therapeutic blood rapamycin levels required for cancer treatment and organ transplant rejection prevention may not be applicable for promoting healthy aging and longevity. Further, patients receiving rapamycin for these conditions have compromised physiological states and may be taking multiple other drugs [21, 22]. Therefore, the PK data from this clinical use population may not effectively translate to gerotherapeutic use in healthy individuals [9, 14]. This is an important consideration in the context of rapamycin, as taking it for geroprotective benefits requires a nuanced approach. Specifically, it must prioritize avoiding adverse events (AEs) to achieve a narrow (likely tissue-specific) geroprotective therapeutic dose range while simultaneously minimizing side effects [23-25].

A small but impactful body of work from Joan Mannick’s lab has provided evidence of safety and tolerability of low-dose rapamycin, and further demonstrates improvements for the aging immune system in a cohort of elderly humans [26, 27]. While this provided important baseline parameters on dosing and dosing schedules for gerotherapeutic applications of rapamycin, many questions remain. Despite these unknowns, recent estimates suggest that over 2,000 people across the USA are currently taking rapamycin off-label [26, 28]. Indeed, we recently described real-world data from a cohort of 333 participants using off-label rapamycin across a wide range of dosages, regimens, and formulations [29]. This study suggested rapamycin can be used safely in normative aging adults over extended periods of time with over one-third of rapamycin users self-reporting benefits in mood, pain, cognition, and fewer moderate to severe acute coronavirus disease 2019 (COVID-19) cases than non-users over the study time period [29]. While this initial evidence is promising, effective validation of rapamycin as a gerotherapeutic requires a deeper exploration of the bioavailability, kinetics, and therapeutic blood rapamycin levels that must be achieved in normative aging individuals for therapeutic effects.

Users of rapamycin off-label as a gerotherapeutic are increasingly turning to its compounded forms, which provide more precise dose tailoring, higher dose capsules for easier administration, easier placebo capsule design for RCTs, and greater affordability [30, 31]. Although rapamycin is categorized as a small molecule, it is relatively large for its class and is lipophilic, resulting in a low solubility in the digestive tract, which may compromise its absorption and bioavailability [20, 32, 33]. This has resulted in broader concerns about the bioavailability of compounded rapamycin [34, 35]. Additionally, individual characteristics (e.g. sex, body mass index (BMI), diet, and genetic polymorphisms) have been shown to influence the bioavailability of rapamycin, but are not yet well understood [36-39]. Further, differing methodologies and quality control practices between compounding pharmacies may result in meaningfully different products [40]. Taken together, skepticism surrounding the use and bioavailability of compounded rapamycin remains, despite its potential advantages.

To begin to address this knowledge gap, we conducted a retrospective study with real-world data on the relative bioavailability and dose-to-blood level linearity of generic and compounded rapamycin by measuring 24-hour blood levels, stratified by dosage, sex, and BMI, in a normative aging cohort. We evaluated inter-individual heterogeneity in bioavailability between different demographics and intra-individual heterogeneity in individuals taking the same dose of rapamycin and escalating doses across different time points. Further, we include a brief review of the literature to place our data in the context of currently available knowledge on rapamycin’s bioavailability in the normative aging population. These efforts aim to contribute to the development of more effective rapamycin longevity protocols, and to inform the design of more rigorous RCTs aimed at validating rapamycin as a putative gerotherapeutic.

## Methods

### Study design

This study collected real-world evidence to better understand and define the relative bioavailability ratio between compounded rapamycin and generic rapamycin, with a specific focus on relative efficacy in increasing rapamycin levels in the blood (blood rapamycin levels). This was a retrospective study, the protocol for which was approved by the Institutional Review Board of the Institute of Regenerative and Cellular Medicine (IRCM, approval number IRCM-2022-352). All participants were provided information about the research being conducted in accordance with Good Clinical Practice (GCP) standards and spoke with a research associate prior to providing informed consent via electronic signature to participate in the research.

### Participants

All participants were healthy, active AgelessRx patients seeking rapamycin treatment (compounded or generic) for its potential longevity and health improvement benefits. Participants who sought rapamycin prescription for longevity support, pain reduction, energy improvement, weight loss, mood improvement, sleep improvement, or oral health improvement were included in the study. To be eligible to receive AgelessRx’s standard rapamycin treatment protocol, participants had to be ≥40 years of age, without a history of uncontrolled disease. All patients treated with rapamycin and opted for at least one blood draw test between May 1, 2023, and December 15, 2023, were included in this retrospective study. After each prescribed dose, patients were given the option to opt in for a blood draw. Those opting in received a survey and requisition for the blood draw.

### Treatment

Those requesting rapamycin and deemed eligible for prescription by the medical team were prescribed compounded or generic rapamycin, based on personal preference. We included 23 participants who were prescribed compounded rapamycin at dosages 5 mg, 10 mg, or 15 mg, beginning at 5 mg/week and titrating up to 15 mg/week, if tolerated without AEs. Further, 44 participants were included who were prescribed generic rapamycin at dosages of 2 mg, 4 mg, 6 mg, or 8 mg. Titration began at 2 mg/week and was titrated up to a target dose of 6 mg/week (option to increase to 8 mg/week if requested by the participant) in 2 mg increments, if needed. The generic (tablets; Dr. Reddy’s Laboratories Limited, Princeton, NJ, USA) and compounded (tablets) rapamycin were dispensed and distributed to participants by Precision Pharmacy (New York, NY, USA). Of important note, as compounded rapamycin is more subject to impurities and inefficacy if vendors do not have adequate quality control and assurance certifications to ensure purity, the compounded rapamycin utilized in our study came from a single pharmacy that was carefully vetted for rigorous quality control and good manufacturing practices (GMP) certifications to limit variability caused by different compounding pharmacy protocols and practices. Careful evaluation of compounded rapamycin sources is critical for appropriate use. Dosing advice for all groups was to take their formulation of rapamycin on the same day of each week, any time of day, with or without meals.

### Assessments

All participants opting in to participate in blood testing for rapamycin levels were sent surveys to complete. The survey asked three questions on rapamycin dosage administered, date and time of administration, and any additional information they wished to share with the AgelessRx research team, in which they were prompted to provide open-ended responses. Participants were instructed to go to the nearest Quest Diagnostics Laboratory location for a blood draw as close to 24 hours after rapamycin administration as possible to measure blood sirolimus (generic name of rapamycin) levels. The research team discussed Quest lab locations with each participant before the start of their rapamycin protocol. A requisition for the ‘Sirolimus assay’ (Test code 36712; Quest Diagnostics) was provided to Quest Diagnostics in advance. This test is designed to measure blood sirolimus/rapamycin levels by liquid chromatography/tandem mass spectrometry. Quest Diagnostics reported the collection date and time with the results of the Sirolimus assay to AgelessRx. The time and date were compared with the participant-completed survey to determine the time between dosage and blood collection.

As the option to participate was sent each time a participant expressed interest in measuring blood rapamycin levels, several interested participants completed additional blood draws to evaluate their Sirolimus assay values while taking the same or higher dosages of rapamycin, based on their titration schedule. Each dose of rapamycin was spaced apart by one week, per the standard rapamycin protocol for healthy aging, ensuring participants’ rapamycin levels were as close to below detectable limits by the Sirolimus assay as possible at the time of the next dosing. Twenty-one participants provided multiple blood draws, which included 15 participants in the compounded group (12 using a different dose, and 3 with the same dose) and 6 participants in the generic group (1 with a different dose, and 5 with the same dose) (**Supplementary Table S1**). To account for the self-selection bias in these groupings, analyses were adjusted accordingly. Notably, cross-sectional analyses were performed using the value obtained after the most recent dose and Sirolimus assay reading (unless otherwise described). We chose this methodology with the assumption that the most recent dose was the dosage determined to be the optimal maintenance dose for the participant and captured the highest doses of rapamycin taken, which is of most clinically relevant interest. Further, as most individuals were taking rapamycin for different periods of time, assessing the most recent dose helped ensure we were uniformly collecting data from individuals taking rapamycin for longer periods of time. For the purposes of this report, we will use the wording blood rapamycin levels when referring to the Sirolimus assay results.

### Statistical analysis

Normality was tested using the Shapiro-Wilk test and visual inspection of Q-Q plots. Generic and compounded groups were compared using Student’s t-test or Welch’s t-test in case of unequal variances for continuous measures, and Chi-squared for categorical measures. Due to the constraints of the real-world retrospective study, data on participants’ baseline levels of rapamycin prior to taking their rapamycin dose was not available, and as such was not included in this study. To test for the presence of bioavailability, one-sample t-tests were performed for each dosage to compare blood rapamycin levels to a hypothesized null value. Similarly, linear mixed-effects models accounting for repeated measures were used to determine whether rapamycin bioavailability differed from zero. Finally, linear mixed-effect models with random intercept per subject were fitted to test for the main effect of protocol (compounded vs. generic) and the interaction effect of protocol by dosage. The models included repeated measures as a within-subject factor and were adjusted for age, sex, and BMI. Statistical analysis was conducted using SPSS 28 (IBM Corp, Armonk, NY, USA), and Python 3.8 using the SciPy and matplotlib packages for data visualization. P-values <0.05 were considered statistically significant.

## Results

### Demographics

This retrospective study evaluated real-world data from 67 participants, of whom 23 received compounded rapamycin and 44 received generic rapamycin (**Supplementary Table S1**). Participants’ age was significantly higher in the compounded group (mean = 61.7 years, SD = 9.1) than in the generic group (mean = 56.8 years, SD = 9.3; t(65) = -2.050, p=0.044; **Table 1**). No significant difference was observed in the mean BMI between the compounded group (mean = 23.8 kg/m^2^, SD = 2.9) and the generic group (mean = 25.1 kg/m^2^, SD = 4.8; t(65) = 1.158, p = 0.251; **Table 1, Supplementary Figure S1**). The compounded group consisted of 73.9% males and in the generic group 59.1% of participants were male (χ**^2^** (1, N = 67) = 1.443, p = 0.230; **Table 1, Supplementary Figure S1**).

**Table 1:** Comparative Demographic Distributions Across Formulation Types.

| | Total | Compounded rapamycin | Generic rapamycin | t(df) or $\chi^2$(df) | p-value |
| --- | --- | --- | --- | --- | --- |
| Age in years, mean (SD) | 58.6 (9.5) | 61.7 (9.1) | 56.8 (9.3) | **-2.050 (65)** | **0.044** |
| BMI (kg/m<sup>2</sup>), mean (SD) | 24.7 (4.3) | 23.8 (2.9) | 25.1 (4.8) | 1.158 (65) | 0.251 |
| Sex, n female (%) | 24 (35.8) | 6 (25.0) | 18 (75.0) | $\chi^2$ (1)= 1.443 | 0.230 |
| Sex, n male (%) | 43 (64.2) | 17 (39.5) | 26 (60.5) |  |  |

BMI: body mass index; SD = standard deviation

In the compounded group, 15 participants had taken two separate blood rapamycin level measurements (65.2%), which in 12 cases were two measurements after a different (increasing) dose (80%) and in 3 cases with the same dose (20%). In the generic group, two separate measurements were available for six participants (13.6%), of which one was with different (increasing) doses (16.7%) and five with the same dose (76.4%). Therefore, 38 data points were available for compounded rapamycin and 50 data points for generic rapamycin (**Supplementary Table S1**).

### Generic and compounded rapamycin result in similar blood rapamycin levels, despite the higher bioavailability of generic rapamycin

In our analyses, we included 1 participant receiving 5 mg, 11 participants receiving 10 mg, and 11 participants receiving 15 mg in the compounded group, while in the generic group, 19 participants received 2 mg, 14 participants received 4 mg, 10 participants received 6 mg, and 1 participant received 8 mg of rapamycin (**Figure 1a**, **Table 2**).

**Figure 1.**
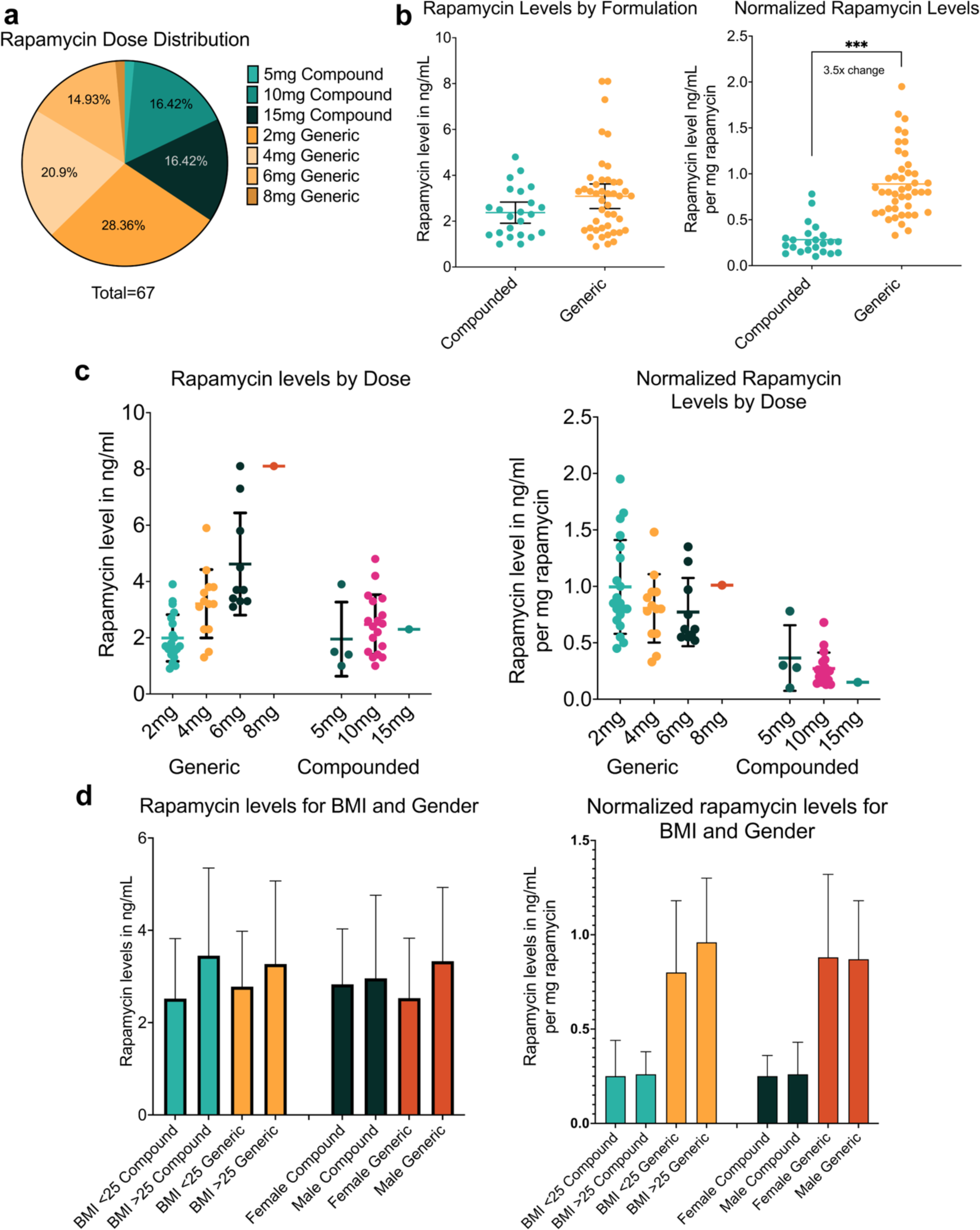
Impact of rapamycin formulation and dose on bioavailability. Quantities of rapamycin (in mg) taken by participants encompassed a moderate span of the gerotherapeutic dosing range **(a)**. Raw values (in ng/ml) of rapamycin concentration in the blood were relatively similar for compounded (mean = 2.93, SD = 1.6) and generic (mean = 3.00, SD = 1.5) formulations across all doses **(b)**; however, normalizing blood rapamycin levels to the amount of rapamycin taken revealed compounded rapamycin (mean = 0.25 ng/mL per 1 mg) is 28.7% less bioavailable per milligram than generic formulations (mean = 0.87 ng/mL per 1 mg; t(32.2) = -9.677, p < 0.001). Stratifying by dose and formulation **(c)** demonstrated bioavailability greater than chance in all instances (all p < 0.001), and no differences in bioavailability could be explained by either BMI or biological sex at birth **(d)** for either raw or normalized blood rapamycin levels (all p non-significant). ***p < 0.001, *BMI: body mass index*.

**Table 2:** Dosage taken and blood rapamycin level of most recent measurements.

|  |  |  |  |  |  |
| --- | --- | --- | --- | --- | --- |
|  | **Total** | **Compounded** | **Generic** | **t(df)** | **p-value** |
| **Most recent measurement** |  |  |  |  |  |
| Dose in mg, mean (SD) | 3.6 (2.3) | 6.4 (0.9) | 2.1 (1.3) | 16.116 (59.1) | **<0.001** |
| Dose, n (%) |  |  |  |  |  |
| 2 mg | 19 (28.4) | - | 19 (43.2) |  |  |
| 4 mg | 14 (20.9) | - | 14 (31.8) |  |  |
| 5 mg | 1 (1.5) | 1 (4.4) | - |  |  |
| 6 mg | 10 (14.9) | - | 10 (22.7) |  |  |
| 8 mg | 1 (1.5) | - | 1 (2.3) |  |  |
| 10 mg | 11 (16.4) | 11 (47.8) | - |  |  |
| 15 mg | 11 (16.4) | 11 (47.8) | - |  |  |
| **Blood levels** |  |  |  |  |  |
| Rapamycin ng/mL, mean (SD) | 2.98 (1.5) | 2.93 (1.6) | 3.00 (1.5) | -0.196 (65.0) | 0.422 |
| Rapamycin ng/mL /1 mg dose, mean (SD) | 0.66 (0.4) | 0.25 (0.2) | 0.87 (0.4) | -9.677 (63.2) | **<0.001** |

*SD = standard deviation*
Note: p-values are derived from Student's t-test, or Welch's t-test for unequal variance.

The mean blood rapamycin concentration in ng/mL in the compounded group was 2.93 (SD = 1.6) and 3.00 (SD = 1.5) in the generic group (t(65) = -0.196, p = 0.422), suggesting similar blood rapamycin levels were achieved across generic and compounded groups despite differences in dosing. Because of this variation in dosages, we also standardized the data by determining the mean blood rapamycin concentration per 1 mg of rapamycin taken. This resulted in a mean rapamycin concentration of 0.25 ng/mL per 1 mg dose for the compounded group, and 0.87 ng/mL per 1 mg dose for the generic group (t(32.2) = -9.677, p < 0.001). This reflects a mean 28.7% bioavailability of compounded rapamycin relative to generic rapamycin in ng/mL / mg rapamycin, suggesting significantly higher rapamycin absorption following generic dosing (**Figure 1b**, **Table 2**). Similar results were obtained when analyzing all data points collected, including multiple measurements within one participant (**Supplementary Table S2**).

As this was a real-world retrospective study, none of the participants included had taken baseline rapamycin blood tests prior to taking their rapamycin dose. However, prior PK data and half-life kinetics with similar doses suggest that blood rapamycin levels would be lower than detectable levels in participants six days after their previous rapamycin dose, making it unlikely that participants previous dose affected their 24-hour measurements due to residual rapamycin in the bloodstream. Therefore, one-sample t-tests were performed comparing evaluable blood rapamycin levels at different dosages to a hypothesized null value to determine if blood rapamycin levels were significantly different from 0. Significant differences were observed at all dose levels (all p<0.001, see **Table 3** and **Supplementary Table S3**), suggesting both compounded and generic rapamycin formulations were bioavailable across the dosing range explored in this study. Specifically, in the compounded rapamycin formulation group, 5 mg rapamycin was taken by one participant resulting in a mean rapamycin level of 3.90 ng/mL, (0.78 ng/mL per mg taken). Eleven participants took 10 mg rapamycin, resulting in a mean rapamycin level of 2.11 ng/mL (SD = 1.1; 0.21 ng/mL per mg taken, SD = 0.1), and another 11 participants took 15 mg rapamycin, resulting in a mean of 3.66 ng/mL (SD = 1.8; 0.25 ng/mL per mg taken, SD = 0.1; **Figure 1c**, **Table 3**). In the generic rapamycin formulation group, 19 participants took 2 mg, resulting in a mean rapamycin level of 2.02 ng/mL (SD = 0.8; 1.01 ng/mL per mg taken, SD = 0.04), 14 participants took 4 mg rapamycin resulting in a mean rapamycin level of 3.24 ng/mL (SD = 1.2; 0.81 ng/mL per mg taken, SD = 0.3), 10 participants took 6 mg rapamycin resulting in a mean rapamycin level of 4.34 ng/mL (SD = 1.8; 0.72 ng/mL per mg taken, SD = 0.3), and 1 participant took 8 mg rapamycin resulting in a mean rapamycin level of 5.0 ng/mL (0.63 ng/mL per mg taken; **Figure 1c**, **Table 3**). Similar results were obtained regardless of whether multiple measurements within one participant were included (**Supplementary Table S3**).

**Table 3.** Bioavailability test of compounded rapamycin at the most recent dose.

|  | Mean blood rapamycin |  |  |  |  |
| --- | --- | --- | --- | --- | --- |
|  | N | levels (SD)<br>in ng/mL | CV | t(df) | p-value |
| Compounded | 23 | 2.93 (1.6) | 0.55 | 8.704 (22) | <0.001 |
| 5 mg | 1 | 3.90 (-) | - | - | - |
| 10 mg | 11 | 2.11 (1.1) | 0.50 | 6.613 (10) | <0.001 |
| 15 mg | 11 | 3.66 (1.8) | 0.49 | 6.790 (10) | <0.001 |
| Compounded /1 mg | 23 | 0.25 (0.2) | 0.64 | 7.783 (22) | <0.001 |
| 5 mg /1 mg | 1 | 0.78 (-) | - | - | - |
| 10 mg /1 mg | 11 | 0.21 (0.1) | 0.52 | 6.613 (10) | <0.001 |
| 15 mg /1 mg | 11 | 0.25 (0.1) | 0.44 | 7.266 (10) | <0.001 |
| Generic, ng/mL | 44 | 3.01 (1.5) | 0.50 | 13.111 (43) | <0.001 |
| 2 mg | 19 | 2.02 (0.8) | 0.42 | 10.467 (18) | <0.001 |
| 4 mg | 14 | 3.24 (1.2) | 0.36 | 10.319 (13) | <0.001 |
| 6 mg | 10 | 4.34 (1.8) | 0.40 | 7.831 (9) | <0.001 |
| 8 mg | 1 | 5.0 (-) | - | - | - |
| Generic /1 mg | 44 | 0.87 (0.4) | 0.43 | 15.847 (43) | <0.001 |
| 2 mg /1 mg | 19 | 1.01 (0.4) | 0.42 | 10.476 (18) | <0.001 |
| 4 mg /1 mg | 14 | 0.81 (0.3) | 0.36 | 10.384 (13) | <0.001 |
| 6 mg /1 mg | 10 | 0.72 (0.3) | 0.40 | 7.870 (9) | <0.001 |
| 8 mg /1 mg | 1 | 0.63 (-) | - | - | - |

CV: coefficient of variation; SD = standard deviation.
Note: p-values derived from one-sample t-tests.

### Rapamycin bioavailability is not significantly influenced by BMI or sex

When analyzing the total group of participants taking either compounded or generic rapamycin, we did not detect significant differences in mean blood rapamycin levels between those with BMI <25 kg/m^2^ and ≥25 kg/m^2^, with compounded achieving mean blood rapamycin levels of 2.52 (SD = 1.3) and 3.45 (SD = 1.9) ng/mL (t(21) = -1.396, p = 0.177) and for generic mean levels were 2.78 (SD = 1.2) and 3.27 (SD = 1.8) ng/mL (t(42) = -1.059, p = 0.296), respectively (**Figure 1d**). The relative blood rapamycin levels for the compounded group were 0.25 (SD = 0.19) and 0.26 (SD = 0.12) and for the generic group 0.80 (SD = 0.38) and 0.96 (0.34) ng/mL per mg taken, for BMI <25 kg/m^2^ and ≥25 kg/m^2^, respectively (t(21) = -0.081, p = 0.936 and t(42) = -1.510, p = 0.139; **Figure 1d**).

Similarly, we did not detect significant differences between sexes with compounded rapamycin resulting in mean blood rapamycin levels of 2.83 (SD = 1.2) and 2.96 (SD = 1.8) (t(21) = -0.160, p = 0.874), and generic resulting in mean levels of 2.53 (SD = 1.3) and 3.33 (SD = 1.6) (t(42) = -1.774, p = 0.083), for female and male participants, respectively (**Figure 1d**). The relative blood rapamycin levels were 0.25 (SD = 0.11) and 0.26 (SD = 0.17) for compounded and 0.88 (SD = 0.44) and 0.87 (SD = 0.31) ng/mL per mg taken for generic, for male and female participants, respectively (t(21) = -0.99, p = 0.848 and t(42) = 0.014, p = 0.989; **Figure 1d**).

Additional evaluations were performed to determine if factors such as age, length of time taking rapamycin, pre-existing health conditions, or other medications taken simultaneously impacted blood rapamycin levels. In all cases, no meaningful relationship was observed (all non-significant, data not shown).

### Rapamycin formulations exhibit a linear dose-to-blood level relationship, but with substantial inter-individual heterogeneity

We determined how the different dosages of compounded and generic rapamycin compared to the measured rapamycin concentration in blood 24 hours after administration. A mixed effect model including all data points (including multiple blood rapamycin level measurements by a single individual) was conducted to test if there were significant differences in dose (mg)-to-rapamycin (ng/mL) blood level relationship between compounded and generic rapamycin. Within both compounded (B = 0.173, SE = 0.071, t(25.6) = 2.442, p = 0.022; 95% CI = 0.027 - 0.319) and generic (B = .697, SE = 0.111, t(38.5) = 6.269, p < 0.001, 95% CI = 0.472 - 0.922) groups, significant associations were observed between dose and blood rapamycin levels (**Figure 2a, Supplementary Table S4.1**). Furthermore, there was a significant interaction effect of dosage by formulation (B = -0.524; SE = 0.130, t(78.6) = -4.026, p < 0.001; 95% CI= - 0.783 - -0.265; **Figure 2a, Supplementary Table S4.2**), indicating the slopes of the dose-to-blood curve were significantly different between compounded and generic rapamycin. Together, these data suggest that while both formulations had linear dose-to-blood level relationships, generic rapamycin elicits a significantly stronger response in blood rapamycin levels as dosage is increased compared to compounded (interaction effect p < 0.001).

**Figure 2.**
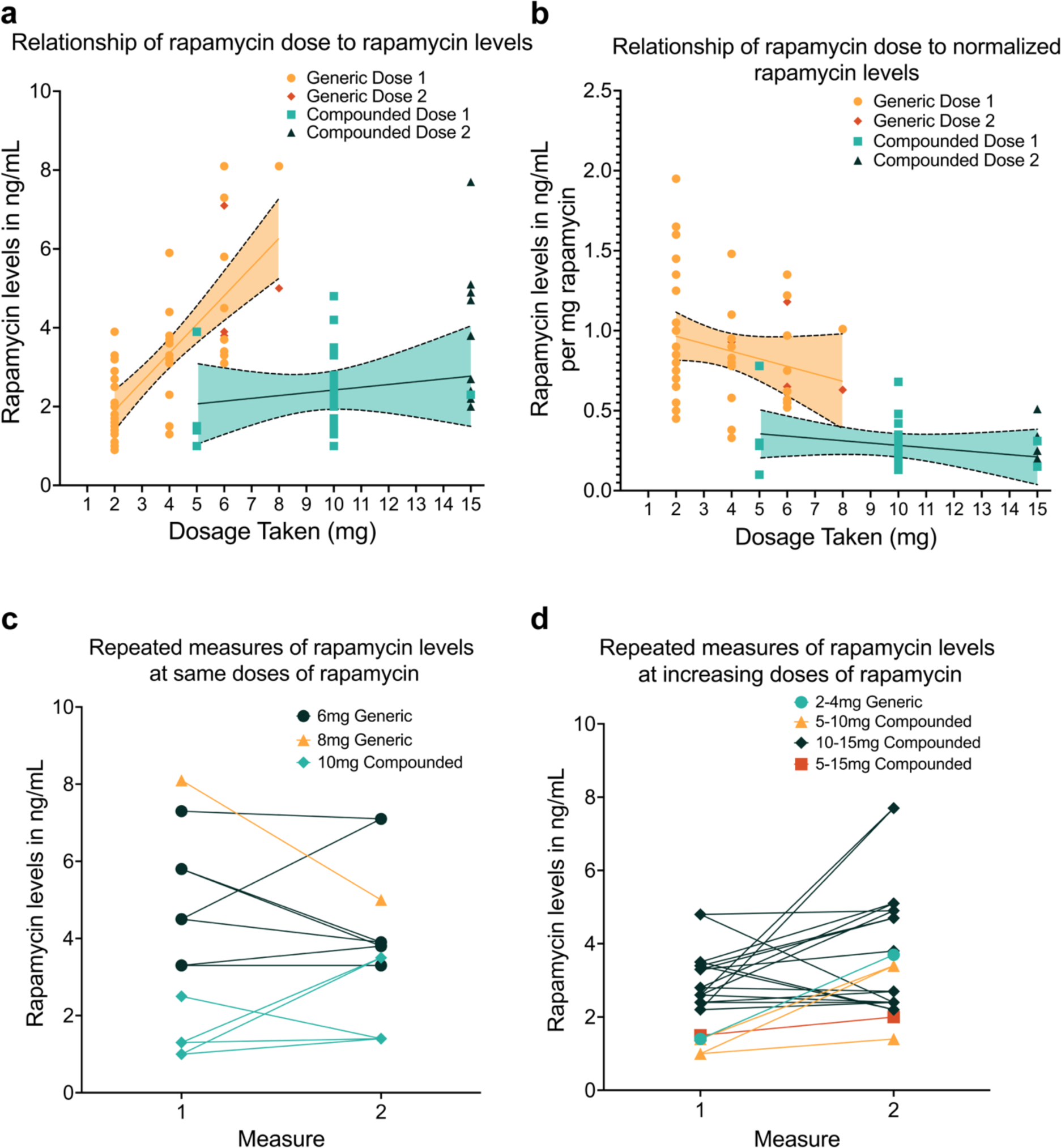
Heterogeneity of dose-to-blood level bioavailability. Compounded and generic formulation groups both demonstrated significant association between dose and blood level (compounded B = .173, SE = 0.071, t(25.6) = 2.442, p = 0.022; 95% CI = 0.027 - .319, generic B = .697, SE = 0.111, t(38.5) = 6.269, p < 0.001, 95% CI = 0.472 - 0.922), with a significant interaction effect (B: -0.524; SE: 0.130, t(78.6) = -4.026, p < 0.001; 95% CI = -0.783 - -0.265) suggesting significantly different increases in bioavailability by formulation across doses **(a)**. Normalized values **(b)** reveal that while more rapamycin was absorbed into the blood per mg delivered in the generic rapamycin group (B = -0.600, SE = 0.320, t(73.6) = -3.619, p < 0.001; 95% CI = -0.930 - -0.270]), bioavailability per mg of rapamycin does not change significantly by formulation as dose increases (interaction non-significant, B = 0.025, SE = 0.028, t(77.9) = 0.911, p = 0.365, 95% CI = -0.030 - 0.081). Despite high inter-individual heterogeneity in bioavailability at a given dose of rapamycin, repeated doses in the same individuals showed consistency in bioavailability for raw and normalized doses **(c)**, and increasing doses in the same individuals tended to increase bioavailability for raw and normalized doses **(e)**, with no meaningful differences between formulations. *CI: confidence interval; SE: standard error*.

While there was a main effect of formulation (B = -0.600, SE = 0.320, t(73.6) = -3.619, p < 0.001; 95% CI = -0.930 - -0.270]) across doses when normalized blood rapamycin levels ((ng/mL)/1 mg) were assessed, the interaction term (dosage by formulation) was non-significant (B = 0.025, SE = 0.028, t(77.9) = 0.911, p = 0.365, 95% CI =-0.030 - 0.081]; **Figure 2b, Supplementary Table S4.3**). This indicates that while more rapamycin was absorbed into the blood per mg delivered in the generic rapamycin group, there was no significant difference between groups in the change in bioavailability per mg rapamycin delivered as the dose increased. There was substantial inter-individual variability in blood rapamycin level concentrations at each dose of compounded and generic rapamycin administered, suggesting differences in bioavailability between people taking the same rapamycin dose independent of formulation (**Figure 2a, b**).

### Rapamycin blood concentration demonstrates variability in response to the same or an escalating dose of rapamycin in an individual

Finally, we assessed the intra-individual heterogeneity of rapamycin blood values in a subset of participants for whom there were multiple measurements with the same dose of rapamycin (compounded, n = 3; generic, n = 5). blood rapamycin levels were similar between the first and second measurement of the same dose for most (though not all) participants, suggesting a relatively stable response (**Figure 2c, Supplementary Figure S2a**). Among participants receiving two differing doses, the second (higher) dose tended to be followed by a higher blood concentration of rapamycin, though some variability in response was again observed (**Figure2d, Supplementary Figure S2b**). No meaningful differences were observed in trends between compounded and generic formulations.

## Discussion and Review

As the geroscience field begins to validate geroprotective agents for their efficacy in improving healthy longevity, rapamycin has garnered particular interest [14]. As little is known about the bioavailability of different formulations of rapamycin in humans, especially in the context of healthy aging, the current study aimed to determine the relative bioavailability of generic and compounded rapamycin by measuring 24-hour blood rapamycin levels in a normative aging cohort to better understand clinical impacts of such in our current and future work. In all participants included in this study, rapamycin could be detected in the blood 24 hours after rapamycin administration. This indicates that both generic and compounded rapamycin are absorbed and bioavailable.

The bioavailability of compounded rapamycin is largely unknown and experts within the longevity field consistently cite concerns with reports of its instability and lack of absorption into the body [41]. Despite these concerns, compounded rapamycin has distinct advantages in cost, accessibility, dosing precision, and placebo creation. Therefore, we aimed to evaluate the bioavailability of compounded rapamycin relative to its generic counterpart using 24-hour blood rapamycin levels normalized to 1 mg of rapamycin.

Our data suggest that compounded rapamycin is bioavailable and has an average 0.287 (28.7%) relative bioavailability to generic rapamycin in (ng/mL) / mg rapamycin. This ratio suggests that the blood rapamycin level response from compounded rapamycin will be approximately 28.7% of blood levels achieved with the same dose of generic rapamycin after 24 hrs. While this relative ratio is important to keep in mind for those utilizing compounded rapamycin, further validation is needed within proper PK studies in normative aging individuals. Further, tracking of blood rapamycin levels over longer periods of time will permit a more complete understanding of the dynamics of the peak concentration, half-life, and other PK parameters that will enable more effective prescribing.

### PK of rapamycin: Dosing for disease vs. healthy aging

The majority of clinical data on rapamycin comes from studies related to its FDA-approved use in patients with organ transplants, cancer, and tuberous sclerosis [42]. Accordingly, the PK of rapamycin has yet to be robustly elucidated in normative aging individuals. Determining the PK in this population is essential to establish optimal dosing regimens to achieve longevity-enhancing effects, particularly as there are likely to be significant physiological differences in the absorption, distribution, metabolism, and excretion (ADME) of rapamycin compared to compromised populations [26].

Rapamycin is a hydrophobic molecule and has been shown to accumulate within the membranes of blood cells (as indicated by a high ratio of whole blood rapamycin levels relative to plasma), particularly red blood cells (RBCs) [43]. This has been suggested to play a major role in observed PK, as it results in bimodal peaks and biphasic clearance. Bimodal peaks are characterized by an initial rapid peak in rapamycin followed by short-term clearance (through cytochrome p450 metabolism and insertion into cellular membranes), and then a second peak in which concentrations more gradually rise again once levels in the blood drop to the point at which the concentration gradient favors release of rapamycin from RBC membranes. This second peak is followed by a more gradual clearance that is thought to give rise to a plateau, or extended tail, of long-duration low blood rapamycin levels and the 40-60 hour terminal half-life observed in organ transplant patients [39, 43-45]. This unique PK behavior may result in the sustained rapamycin activity shown to drive the therapeutic benefits experienced with low-dose, intermittent (e.g. weekly) dosing. While additional factors such as the number of RBCs and their functional characteristics may influence rapamycin PK and effectiveness, further study is required to fully elucidate this relationship [20, 46]. Regardless, the complexities of rapamycin PK may help explain how the lower 24-hour blood rapamycin levels observed in this study can have sustained effects that drive therapeutic benefits.

Data on therapeutic levels of rapamycin in the literature are largely focussed on its use in individuals with disease. However, these data are limited by the compromised physiology of the individual, daily and higher dosing required for efficacy, and the presence of other drugs that are co-administered (such as chemotherapeutics, steroids, and other immunosuppressants) [30]. Though little is known about the therapeutic blood rapamycin levels and safety over longer periods of time in normative aging individuals taking rapamycin for healthy aging, recent research suggests that low blood rapamycin levels are sufficient for exerting functional benefits on multiple cell types and organ systems [14, 22, 46]. For example, *in vitro* tumor angiogenesis was inhibited in human lymphangioma cells cultured at rapamycin concentrations as low as 1 ng/mL [47].

In humans, higher doses (for example, 3 mg/day) of mTOR inhibitors inhibit T cell function and are used to suppress the immune response to avoid immune-mediated organ transplant rejection (at target trough blood rapamycin levels of 5-15 ng/mL) [14]. In contrast to this immunosuppressive dose, 6-fold lower doses have been associated with mitigation of age-related decline in immune function [14]. While further investigation into the optimally effective dose and duration of rapamycin treatment is undoubtedly required, these studies suggest significant promise for low-dose rapamycin and its derivatives in positively impacting healthy aging.

The 24-hour rapamycin blood concentration for most participants in our study was between 1-4 ng/mL (and as high as 8.1 ng/ml), regardless of the use of compounded or generic formulation of rapamycin. This aligns with the therapeutic blood range associated with immune benefits in normative aging cohorts ([27] & correspondence with Joan Mannick). While these 24-hour blood rapamycin levels likely lead to low blood rapamycin levels in the days prior to the next rapamycin dosage, *in vitro* studies suggest that rapamycin can inhibit mTOR at concentrations of as little as 1 ng/mL, suggesting it can effectively induce cellular effects even at these low levels [48].

The 24-hour time point measurement in this real-world study was chosen for participant convenience based on their dosing time as recommended by the AgelessRx medical team. Although data suggest that 24-hour time point measurements correlate with its bioavailability and therapeutic dose, it is unknown whether measuring 24-hour blood rapamycin levels is indicative of whether therapeutic bioavailability is attained as it represents only a cross-sectional snapshot of rapamycin’s bioavailability [49, 50]. It also remains unknown how long and at what level rapamycin should be present in the body for longevity-promoting effects [25]. Measuring Cmax (peak concentrations) of rapamycin may be a better indicator of bioavailability, as it reflects the total amount of rapamycin in the blood before clearance begins [46, 51]. However, measuring peak blood rapamycin levels within our participants presented several challenges. Most importantly, peak blood rapamycin levels vary greatly as they occur extremely fast and there can be significant differences in measured blood rapamycin levels within a short period of time [46, 52]. Conversely, several PK studies suggest trough concentrations correlate with the efficacy and safety of rapamycin, as it has a long tail and thus represents the largest exposure time of organs to rapamycin [52, 53]. While it was beyond the scope of the current study to explore this further, future studies measuring blood rapamycin levels at peak, trough, and intermediate time points are undoubtedly required to more fully understand the best time points at which to measure rapamycin concentrations to correlate with clinical outcomes.

### Factors affecting rapamycin PK in a normative aging cohort

The annotated rapamycin half-life in the literature for compromised populations is 60-80 hours [48, 51]. However, the blood rapamycin levels detected after 24 hours in the current study (most below 4 ng/mL and none above 8 ng/mL) suggest the half-life might be different in normative aging populations. This is consistent with a previous study in which 12 healthy volunteers were given a single 6 mg oral dose of generic rapamycin resulting in plasma concentrations after 24 hours of 4.47 ± 0.57 ng/mL (range: 2.90 – 7.20 ng/mL) [54]. Again, further study will be required to more completely understand these effects, and to determine if specific health factors influence the half-life of rapamycin predictably enough to be considered in therapeutic strategies moving forward.

Several additional factors may influence rapamycin bioavailability, such as microbial interactions, plasma protein binding, hydrophobicity, and interactions with cellular membranes, and an individual’s genetics, diet, underlying physiology, and potentially other lifestyle factors [36, 39]. High-fat meals, grapefruit juice, and certain drugs (e.g. ketoconazole) have been shown to impact rapamycin’s bioavailability by up to 350% in some instances [37, 38]. Grapefruit juice in particular was shown to cause a longer peak and higher sustained levels, which may impact adverse events, while curcumin has been shown to substantially decrease bioavailability (>75%) in animal models [38, 55]. We did not observe any significant relationship of rapamycin bioavailability with sex, BMI, age, pre-existing health conditions, or other medications taken simultaneously. Importantly, we did not detect any relationship between length of time taking rapamycin and bioavailability either, indicating a lack of desensitization from taking the drug over extended periods of time. Future studies will benefit from exploring each of these in greater depth to increase the field’s understanding of how to most effectively utilize rapamycin for longevity medicine.

### Inter-individual heterogeneity in response to rapamycin: Importance of measuring blood rapamycin levels in every patient

As the field evaluates the optimal dosage and regimen to utilize rapamycin for healthy aging, it is essential to understand heterogeneity in response to the same dose of rapamycin between normative aging individuals. Our study suggests significant inter-individual heterogeneity in the relative bioavailability of both compounded and generic rapamycin with a 2-6x difference in blood rapamycin levels in those taking the same dose. Importantly, intra-individual heterogeneity in bioavailability for the same dose of rapamycin showed a high test-retest reliability, suggesting consistent blood rapamycin level results for individuals taking the same dose over time. This is important for the requisite safety and reliability of rapamycin with the long-duration regimens required for aging applications, but also highlights the importance of evaluating blood rapamycin levels in every individual taking rapamycin, regardless of its formulation, to accurately evaluate effectiveness and any AEs.

Inter-individual rapamycin bioavailability variability explained by sex-specific effects have been mixed in preclinical studies to date. For example, Interventions Testing Program (ITP) studies provided mouse chow with 44 ppm rapamycin to UM-HET3 mice for one hour after an overnight fast and observed that blood rapamycin levels were higher in female mice than in males (80 ng/mL and 23 ng/mL, respectively), corresponding with greater lifespan benefits [56]. However, the opposite effect was observed in non-human primates. After dosing with rapamycin (∼ 1 mg/kg body weight per day, 5 days per week), trough concentrations of rapamycin were significantly lower in females compared to male marmosets (4.4 ng/mL and 8.4 ng/mL, respectively). Despite this pharmacological dissimilarity, few differences in basic morphology or hematological markers of blood cell counts, metabolism, or inflammation were observed between male and female marmosets after nine months of treatment [53].

Importantly, limited data is available on the sex-specific availability of rapamycin in humans, and even less in healthy populations. We are the first to report sex-specific rapamycin bioavailability in a normative aging population taking rapamycin for longevity. The current study detected no statistically significant differences in rapamycin bioavailability based on sex or BMI across compounded and generic doses at the 24-hour time point; however, there was a trend (p= 0.083) of higher bioavailability in male participants taking generic rapamycin that should be followed up in future studies. These initial human data imply that any differences in rapamycin effectiveness between sexes (such as those seen in preclinical lifespan studies) may be related to other sex-specific factors involved in rapamycin pharmacodynamics or potentially in rates of rapamycin clearance beyond 24 hours.

### Titrating to longevity dose: linear dose-to-blood rapamycin level relationships

A linear relationship between dose and blood rapamycin levels has previously been shown in studies in disease populations taking rapamycin [57]. This is an important factor for titrating to the appropriate longevity dose of rapamycin, as it makes predictive dosing more feasible. The current study observed the expected linear relationship of rapamycin concentration in ng/mL in the blood with increasing doses of rapamycin administered. However, likely due to differences in the bioavailability ratio between generic and compounded formulations of rapamycin, comparable maximum concentrations were not observed. While a slower increase in blood rapamycin levels with compounded rapamycin could be beneficial as it pertains to the minimization of AEs (immunosuppression and metabolic dysfunction), further study into the PK of rapamycin and the optimal longevity-therapeutic dose is undoubtedly warranted to more comprehensively understand the benefits and limitations of this formulation approach [26].

Previous work suggests that lower (and by extension more granularly dose-controllable) rapamycin doses may have significant longevity benefits [14, 22]. For example, a study conducted by Mannick *et al*. in normative aging elderly individuals demonstrated that participants taking 0.5 mg/day of everolimus (an mTORc1 inhibitor similar to rapamycin) for 6 weeks had a 40% mTOR inhibition with a Cmax of <3 ng/mL and Cmin <1 ng/mL ([27, 58] & correspondence with Joan Mannick). Further, regimens of 0.5 mg/day or 5 mg/week both showed efficacy in enhancing antibody titers in response to influenza vaccination, with no differences detected in AEs, as opposed to 20 mg/week dosing [27]. This suggests that lower blood rapamycin levels present over several days with troughs as low as 1 ng/mL may be sufficient for immune-related benefits. However, these results must be taken in context, as everolimus has higher absorption and bioavailability than rapamycin (sirolimus), but a shorter half-life and faster elimination [59].

Importantly, when graphing the blood rapamycin levels detected across increasing doses, our work has shown that the slopes of the curves between the generic and compounded rapamycin were significantly different across doses. Generic rapamycin had a steeper slope for the dose-to-blood level graph than compounded; however, normalized (to 1 mg rapamycin taken) measures also demonstrated a non-significant decline with increasing dose, suggesting bioavailability saturation. This suggests that higher concentrations of compounded rapamycin may be required to achieve similar bioavailability saturation. However, this is balanced by the more consistent bioavailability of compounded rapamycin compared to generic, suggesting compounded formulations are likely more predictable in effect and AEs, and permit options for AE mitigation through smaller, more confined distribution of blood rapamycin levels.

Finally, because we included data from participants taking multiple blood rapamycin level measurements, it afforded us the opportunity to evaluate the effects of escalating doses of rapamycin in a single individual across different time points. The second (higher) dose tended to be followed by a higher blood concentration of rapamycin, though there was noteworthy inter-individual heterogeneity in response. These findings underscore the importance of measuring blood rapamycin levels upon initial prescription and during dose titration. Given the observed variability in PK, personalized dosing strategies may be necessary to achieve optimal therapeutic outcomes for each patient.

### Conclusion and focus for future research

In conclusion, this study represents the largest investigation to date of real-world evidence on the relative bioavailability of generic and compounded rapamycin in a healthy, normative aging cohort. Our findings suggest that therapeutic blood concentrations can be obtained from both formulations; however, the generic formulation demonstrated approximately 3.5x more potency than compounded. A linear dose-response was observed for both formulations; however, the notably more gradual slope of compounded rapamycin increase suggests that it may be utilized for more specific and granular dosing. This may be an important advantage of compounded rapamycin, as the high rates of inter-individual (but not intra-individual) heterogeneity in response to both formulations indicate dosing should be tailored to each individual and monitored regularly for maximal efficacy.

In keeping with the reduced potency of compounded rapamycin, we observed saturation of bioavailability only with the generic formulation, though average availability of both formulas ranged from 1-4 ng/ml (and as high as 8.1 ng/ml). As the maximal compounded dose utilized in this study was only 15 mg (equivalent to approximately 4.3 mg generic), we likely did not reach adequate dosing range to achieve bioavailability saturation with the compounded formulation in this study. This will be an important consideration for future studies, particularly those exploring greater detail on dosing range, PK, optimal dosing intervals, and ADME of both generic and compound formulations of rapamycin.

Surprisingly, independent of formulation type, we did not observe any significant relationship of rapamycin bioavailability with sex, BMI, age, length of time taking rapamycin, pre-existing health conditions, or other medications taken simultaneously. This suggests that factors such as metabolism, diet, or genetics may significantly impact bioavailability, which should be explored in depth in future studies. Given the pronounced inter-individual (but not intra-individual) heterogeneity of rapamycin in blood after 24 hrs in both formulation groups, we would recommend routine measurement of blood rapamycin levels at standardized time points until a biological basis of heterogeneity and any associated AE changes is more fully understood. Measuring blood rapamycin levels at standardized time points will allow the geroscience field to derive more valuable insights into the correlation of blood rapamycin levels with benefits and AEs.

Taken together, the results of this study suggest that converging on the optimal formulation of rapamycin to utilize (compounded or generic) depends on the delicate balance of the nuances of bioavailability with the practicalities of treatment. We highlight that while compounded rapamycin had a reliable bioavailability and performed well in comparison to generic rapamycin, generic rapamycin demonstrated a stronger dose-to-bioavailability relationship and reached blood saturation at lower doses. We suggest that compounded rapamycin may have practical advantages in clinical practice, including greater accessibility, tailoring of personalized dosages, and the potential of increased adherence (due to cost savings and reduced number of tablets). However, it should be emphasized that due diligence regarding the quality of compounded rapamycin sources is critical for accurate clinical understanding. Our findings provide insights that open the door for future studies to more extensively measure blood rapamycin levels achieved in the context of clinical studies, so that we can continue to build our understanding of rapamycin in promoting longevity.

## Data Availability

All data produced in the present study are available upon reasonable request to the authors

## Statements and Declarations

### Competing interests

GH, VL, AN, MM, BV, CT, SLM, AI, and SZ are employees and shareholders of AgelessRx. MW and JH have received financial compensation from AgelessRx for their contributions. KK is an employee and shareholder of Venture Plant AB and Rapamycin Longevity Lab, which has received a donation from AgelessRx for research coordination in partnership with Ora Biomedical.

### Author contributions

Virginia Lee, Andy Nyquist, Anar Isman, and Sajad Zalzala designed and implemented the study. Girish Harinath, Jesper Hagemeier, Brandon Verkennes, Colleen Tacubao, and Stefanie L Morgan performed data analysis. Girish Harinath, Virginia Lee, Andy Nyquist, Mauricio Moel, Maartje Wouters, Krister Kauppi, Stefanie L Morgan, Anar Isman, and Sajad Zalzala wrote and edited the manuscript. All work was supervised by Stefanie L Morgan, Anar Isman, and Sajad Zalzala.

## Acknowledgements

The authors would like to thank the participants who took part in this study. Financial support, administrative support, and article publishing charges were provided by AgelessRx.

## Notes

### Clinical Trial

NCT06550271

### Funding Statement

This study was funded by AgelessRx.

### Author Declarations

The Institutional Review Board of the Institute of Regenerative and Cellular Medicine (IRCM, approval number IRCM-2022-352) gave ethical approval for this work.

## References

1. Steverson M. Ageing and health. World Health Organization. 2022. https://www.who.int/news-room/fact-sheets/detail/ageing-and-health. Accessed August 2024.

2. Guo J, Huang X, Dou L, Yan M, Shen T, Tang W, et al. Aging and aging-related diseases: from molecular mechanisms to interventions and treatments. Signal Transduct Target Ther. 2022;7(1):391; 10.1038/s41392-022-01251-0.

3. Tian YE, Cropley V, Maier AB, Lautenschlager NT, Breakspear M, Zalesky A. Heterogeneous aging across multiple organ systems and prediction of chronic disease and mortality. Nat Med. 2023;29(5):1221–31; 10.1038/s41591-023-02296-6.

4. Kehler DS. Age-related disease burden as a measure of population ageing. Lancet Public Health. 2019;4(3):e123–e4; 10.1016/S2468-2667(19)30026-X.

5. Centers for Disease Control and Prevention. Trends in aging--United States and worldwide. MMWR Morb Mortal Wkly Rep. 2003;52(6):101-6;

6. Rolland Y, Sierra F, Ferrucci L, Barzilai N, De Cabo R, Mannick J, et al. Challenges in developing geroscience trials. Nat Commun. 2023;14(1):5038; 10.1038/s41467-023-39786-7.

7. Kulkarni AS, Aleksic S, Berger DM, Sierra F, Kuchel GA, Barzilai N. Geroscience-guided repurposing of FDA-approved drugs to target aging: a proposed process and prioritization. Aging Cell. 2022;21(4):e13596; 10.1111/acel.13596.

8. Le Couteur DG, Barzilai N. New horizons in life extension, healthspan extension and exceptional longevity. Age Ageing. 2022;51(8); 10.1093/ageing/afac156.

9. Blagosklonny MV. Towards disease-oriented dosing of rapamycin for longevity: does aging exist or only age-related diseases? Aging (Albany NY). 2023;15(14):6632–40; 10.18632/aging.204920.

10. Sabatini DM. Twenty-five years of mTOR: Uncovering the link from nutrients to growth. Proc Natl Acad Sci U S A. 2017;114(45):11818–25; 10.1073/pnas.1716173114.

11. Eltschinger S, Loewith R. TOR complexes and the maintenance of cellular homeostasis. Trends Cell Biol. 2016;26(2):148–59; 10.1016/j.tcb.2015.10.003.

12. Papadopoli D, Boulay K, Kazak L, Pollak M, Mallette FA, Topisirovic I, et al. mTOR as a central regulator of lifespan and aging. F1000Res. 2019;8; 10.12688/f1000research.17196.1.

13. Zhang Y, Zhang J, Wang S. The role of rapamycin in healthspan extension via the delay of organ aging. Ageing Res Rev. 2021;70:101376; 10.1016/j.arr.2021.101376.

14. Mannick JB, Lamming DW. Targeting the biology of aging with mTOR inhibitors. Nat Aging. 2023;3(6):642–60; 10.1038/s43587-023-00416-y.

15. Urfer SR, Kaeberlein TL, Mailheau S, Bergman PJ, Creevy KE, Promislow DEL, et al. A randomized controlled trial to establish effects of short-term rapamycin treatment in 24 middle-aged companion dogs. Geroscience. 2017;39(2):117–27; 10.1007/s11357-017-9972-z.

16. Toutain PL, Ferran A, Bousquet-Melou A. Species differences in pharmacokinetics and pharmacodynamics. Handb Exp Pharmacol. 2010(199):19–48; 10.1007/978-3-642-10324-7_2.

17. Augustine JJ, Bodziak KA, Hricik DE. Use of sirolimus in solid organ transplantation. Drugs. 2007;67(3):369–91; 10.2165/00003495-200767030-00004.

18. Gallant-Haidner HL, Trepanier DJ, Freitag DG, Yatscoff RW. Pharmacokinetics and metabolism of sirolimus. Ther Drug Monit. 2000;22(1):31-5; 10.1097/00007691-200002000-00006.

19. Hartinger JM, Rysanek P, Slanar O, Sima M. Pharmacokinetic principles of dose adjustment of mTOR inhibitors in solid organ transplanted patients. J Clin Pharm Ther. 2022;47(9):1362–7; 10.1111/jcpt.13753.

20. Trepanier DJ, Gallant H, Legatt DF, Yatscoff RW. Rapamycin: distribution, pharmacokinetics and therapeutic range investigations: an update. Clin Biochem. 1998;31(5):345–51; 10.1016/s0009-9120(98)00048-4.

21. Grogan S, Preuss CV. Pharmacokinetics. StatPearls. Treasure Island (FL)2024.

22. Lee DJW, Hodzic Kuerec A, Maier AB. Targeting ageing with rapamycin and its derivatives in humans: a systematic review. Lancet Healthy Longev. 2024;5(2):e152–e62; 10.1016/S2666-7568(23)00258-1.

23. Blagosklonny MV. Rapamycin for longevity: opinion article. Aging (Albany NY). 2019;11(19):8048–67; 10.18632/aging.102355.

24. Johnson SC, Kaeberlein M. Rapamycin in aging and disease: maximizing efficacy while minimizing side effects. Oncotarget. 2016;7(29):44876–8; 10.18632/oncotarget.10381.

25. Kaeberlein M. Rapamycin and ageing: when, for how long, and how much? J Genet Genomics. 2014;41(9):459–63; 10.1016/j.jgg.2014.06.009.

26. Konopka AR, Lamming DW, Investigators RP, Investigators E. Blazing a trail for the clinical use of rapamycin as a geroprotecTOR. Geroscience. 2023;45(5):2769–83; 10.1007/s11357-023-00935-x.

27. Mannick JB, Del Giudice G, Lattanzi M, Valiante NM, Praestgaard J, Huang B, et al. mTOR inhibition improves immune function in the elderly. Sci Transl Med. 2014;6(268):268ra179; 10.1126/scitranslmed.3009892.

28. Gilbert D. How a cheap, generic drug became a darling of longevity enthusiasts. The Washington Post [Internet]. 2024 [cited 2024 March]. Available from: https://www.washingtonpost.com/business/2024/03/15/rapamycin-longevity-drug/.

29. Kaeberlein TL, Green AS, Haddad G, Hudson J, Isman A, Nyquist A, et al. Evaluation of off-label rapamycin use to promote healthspan in 333 adults. Geroscience. 2023;45(5):2757–68; 10.1007/s11357-023-00818-1.

30. Lisi DM. Pros and cons of pharmacy compounding. US Pharmacist. 2021. https://www.uspharmacist.com/article/pros-and-cons-of-pharmacy-compounding. Accessed August 2024.

31. Carvalho M, Almeida IF. The role of pharmaceutical compounding in promoting medication adherence. Pharmaceuticals (Basel). 2022;15(9); 10.3390/ph15091091.

32. Shi Y, Jiao C, Lu X, Nie Y, Li X, Han D. Rapamycin nanoparticles improves drug bioavailability in PLAM treatment by interstitial injection. Orphanet J Rare Dis. 2022;17(1):349; 10.1186/s13023-022-02511-6.

33. Kuerec AH, Maier AB. Why is rapamycin not a rapalog? Gerontology. 2023;69(6):657–9; 10.1159/000528985.

34. Sharp ZD, Strong R. Rapamycin, the only drug that has been consistently demonstrated to increase mammalian longevity. An update. Exp Gerontol. 2023;176:112166; 10.1016/j.exger.2023.112166.

35. Ehninger D, Neff F, Xie K. Longevity, aging and rapamycin. Cell Mol Life Sci. 2014;71(22):4325–46; 10.1007/s00018-014-1677-1.

36. Stielow M, Witczynska A, Kubryn N, Fijalkowski L, Nowaczyk J, Nowaczyk A. The bioavailability of drugs-The current state of knowledge. Molecules. 2023;28(24); 10.3390/molecules28248038.

37. Zimmerman JJ, Ferron GM, Lim HK, Parker V. The effect of a high-fat meal on the oral bioavailability of the immunosuppressant sirolimus (rapamycin). J Clin Pharmacol. 1999;39(11):1155–61;

38. Cohen EE, Wu K, Hartford C, Kocherginsky M, Eaton KN, Zha Y, et al. Phase I studies of sirolimus alone or in combination with pharmacokinetic modulators in advanced cancer patients. Clin Cancer Res. 2012;18(17):4785-93; 10.1158/1078-0432.CCR-12-0110.

39. Emoto C, Fukuda T, Cox S, Christians U, Vinks AA. Development of a physiologically-based pharmacokinetic model for Sirolimus: Predicting bioavailability based on intestinal CYP3A content. CPT Pharmacometrics Syst Pharmacol. 2013;2(7):e59; 10.1038/psp.2013.33.

40. Gudeman J, Jozwiakowski M, Chollet J, Randell M. Potential risks of pharmacy compounding. Drugs R D. 2013;13(1):1–8; 10.1007/s40268-013-0005-9.

41. Attia P. #272 ‒ Rapamycin: potential longevity benefits, surge in popularity, unanswered questions, and more | David Sabatini, MD, PhD and Matt Kaeberlein, PhD [Internet]; 2023. Podcast. Available from: https://peterattiamd.com/davidsabatini-mattkaeberlein/

42. Mohamed MA, Elkhateeb WA, Daba GM. Rapamycin golden jubilee and still the miraculous drug: a potent immunosuppressant, antitumor, rejuvenative agent, and potential contributor in COVID-19 treatment. Bioresour Bioprocess. 2022;9(1):65; 10.1186/s40643-022-00554-y.

43. Yanez JA, Forrest ML, Ohgami Y, Kwon GS, Davies NM. Pharmacometrics and delivery of novel nanoformulated PEG-b-poly(epsilon-caprolactone) micelles of rapamycin. Cancer Chemother Pharmacol. 2008;61(1):133–44; 10.1007/s00280-007-0458-z.

44. Zhou Q, Doherty J, Akk A, Springer LE, Fan P, Spasojevic I, et al. Safety profile of rapamycin perfluorocarbon anoparticles for preventing cisplatin-induced kidney injury. Nanomaterials (Basel). 2022;12(3); 10.3390/nano12030336.

45. Mahalati K, Kahan BD. Clinical pharmacokinetics of sirolimus. Clin Pharmacokinet. 2001;40(8):573–85; 10.2165/00003088-200140080-00002.

46. Evans JB, Morrison AJ, Javors MA, Lopez-Cruzan M, Promislow DEL, Kaeberlein M, et al. Pharmacokinetics of long-term low-dose oral rapamycin in four healthy middle-aged companion dogs. bioRxiv. 2021:2021.01.20.427425; 10.1101/2021.01.20.427425.

47. Huber S, Bruns CJ, Schmid G, Hermann PC, Conrad C, Niess H, et al. Inhibition of the mammalian target of rapamycin impedes lymphangiogenesis. Kidney Int. 2007;71(8):771–7; 10.1038/sj.ki.5002112.

48. Marx SO, Jayaraman T, Go LO, Marks AR. Rapamycin-FKBP inhibits cell cycle regulators of proliferation in vascular smooth muscle cells. Circ Res. 1995;76(3):412–7; 10.1161/01.res.76.3.412.

49. Leelahavanichkul A, Areepium N, Vadcharavivad S, Praditpornsilpa K, Avihingsanon Y, Karnjanabuchmd T, et al. Pharmacokinetics of sirolimus in Thai healthy volunteers. J Med Assoc Thai. 2005;88 Suppl 4:S157–62;

50. Woodrum C, Nobil A, Dabora SL. Comparison of three rapamycin dosing schedules in A/J Tsc2+/- mice and improved survival with angiogenesis inhibitor or asparaginase treatment in mice with subcutaneous tuberous sclerosis related tumors. J Transl Med. 2010;8:14; 10.1186/1479-5876-8-14.

51. Leung LY, Lim HK, Abell MW, Zimmerman JJ. Pharmacokinetics and metabolic disposition of sirolimus in healthy male volunteers after a single oral dose. Ther Drug Monit. 2006;28(1):51–61; 10.1097/01.ftd.0000179838.33020.34.

52. QuestDiagnostics. Sirolimus (Rapamycin). QuestDiagnostics. https://testdirectory.questdiagnostics.com/test/test-guides/TS_Sirolimus_Rapamycin/sirolimus-rapamycin?gf=y. Accessed August 2024.

53. Sills AM, Artavia JM, DeRosa BD, Ross CN, Salmon AB. Long-term treatment with the mTOR inhibitor rapamycin has minor effect on clinical laboratory markers in middle-aged marmosets. Am J Primatol. 2019;81(2):e22927; 10.1002/ajp.22927.

54. Brattstrom C, Sawe J, Jansson B, Lonnebo A, Nordin J, Zimmerman JJ, et al. Pharmacokinetics and safety of single oral doses of sirolimus (rapamycin) in healthy male volunteers. Ther Drug Monit. 2000;22(5):537–44; 10.1097/00007691-200010000-00006.

55. Hsieh YW, Huang CY, Yang SY, Peng YH, Yu CP, Chao PD, et al. Oral intake of curcumin markedly activated CYP 3A4: in vivo and ex-vivo studies. Sci Rep. 2014;4:6587; 10.1038/srep06587.

56. Miller RA, Harrison DE, Astle CM, Fernandez E, Flurkey K, Han M, et al. Rapamycin-mediated lifespan increase in mice is dose and sex dependent and metabolically distinct from dietary restriction. Aging Cell. 2014;13(3):468–77; 10.1111/acel.12194.

57. MacDonald A, Scarola J, Burke JT, Zimmerman JJ. Clinical pharmacokinetics and therapeutic drug monitoring of sirolimus. Clin Ther. 2000;22 Suppl B:B101-21; 10.1016/s0149-2918(00)89027-x.

58. Kauppi K. Joan Mannick on Rapamycin Longevity Series | Turning down mTOR to young levels may be good for aging [Internet]; 2024. Podcast. Available from: https://www.youtube.com/watch?v=f1b4sxLQD08&t=120s

59. Klawitter J, Nashan B, Christians U. Everolimus and sirolimus in transplantation-related but different. Expert Opin Drug Saf. 2015;14(7):1055–70; 10.1517/14740338.2015.1040388.

