## Supplementary Tables for "The bioavailability of compounded and generic rapamycin in normative aging individuals: A retrospective study and review with clinical implications"

**Supplementary Material**

| **Supplementary Table S1: Overview of participants and number of measurements** | | | |
| --- | --- | --- | --- |
|  | Total n (%) | Compounded n (%) | Generic n (%) |
| **Number of participants** | 67 (100.0) | 23 (100.0) | 44 (100.0) |
| Participants with two    measurements | 21 (31.3) | 15 (65.2) | 6 (13.6) |
| Participants with one    measurement | 46 (68.7) | 8 (34.8) | 38 (86.4) |
| **Participants with two measurements** | 21 (100.0) | 15 (100.0) | 6 (100.0) |
| Measurements with    different doses | 13 (61.9) | 12 (80.0) | 1 (16.7) |
| Measurements with    the same dose | 8 (38.1) | 3 (20.0) | 5 (83.3) |
| **Unique data points^*^** | 88 (100.0) | 38 (100.0) | 50 (100.0) |
| Data points with two    measurements | 42 (47.7) | 30 (78.9) | 12 (24.0) |
| Data points with one    measurement | 46 (52.3) | 8 (21.1) | 38 (76.0) |

*^*^Unique data points reflect the total number of measurements*

| **Supplemental Table S2: Dosage taken and blood rapamycin level of all measurements** | | | | | |
| --- | --- | --- | --- | --- | --- |
|  | **Total** | **Compounded** | **Generic** | **t(df)** | **p-value** |
| **All measurements** |  |  |  |  |  |
| Dose in mg, mean (SD) | 6.94 (3.0) | 10.92 (3.0) | 3.92 (1.9) | **9.142 (75.5)** | **<0.001** |
| Dose, n (%) |  |  |  |  |  |
| 2 mg | 20 (22.7) | - | 20 (40.0) |  |  |
| 4 mg | 14 (15.9) | - | 14 (28.0) |  |  |
| 5 mg | 4 (4.5) | 4 (10.5) | - |  |  |
| 6 mg | 14 (15.9) | - | 14 (28.0) |  |  |
| 8 mg | 2 (2.3) | - | 2 (4.0) |  |  |
| 10 mg | 23 (26.1) | 23 (60.5) | - |  |  |
| 15 mg | 11 (12.5) | 11 (28.9) | - |  |  |
| **Blood levels** |  |  |  |  |  |
| Rapamycin ng/mL, mean (SD)^†^ | 3.03 (0.6) | 2.73 (1.4) | 3.25 (1.8) | -1.565 (84.0) | 0.121 |
| Rapamycin ng/mL /1 mg dose, mean (SD) | 0.61 (0.6) | 0.27 (0.2) | 0.87 (0.4) | **-5.130 (66.6)** | **<0.001** |

*df: degrees of freedom; SD = standard deviation*

Note: p-values were derived from linear mixed-effects models with random intercept, accounting for repeated measures.

| **Supplementary Table S3.** Bioavailability test of compounded rapamycin including all data points | | | | | |
| --- | --- | --- | --- | --- | --- |
|  | **n** | **Mean blood rapamycin levels (SD)** in ng/mL | **CV** | **t(df)** | **p-value** |
| Compounded | 38 | 2.73 (1.4) | 0.52 | 10.607 (22.5) | **<0.001** |
| 5 mg | 4 | 1.95 (1.3) | 0.68 | 2.959 (3.0) | 0.060 |
| 10 mg | 23 | 2.42 (1.1) | 0.43 | 11.113 (15.4) | **<0.001** |
| 15 mg | 11 | 3.66 (1.8) | 0.49 | 6.790 (10.0) | **<0.001** |
| Compounded / 1 mg | 38 | 0.27 (0.2) | 0.56 | 9.425 (16.6) | **<0.001** |
| 5 mg /1 mg | 4 | 0.37 (0.3) | 0.78 | 2.509 (3.0) | 0.087 |
| 10 mg /1 mg | 23 | 0.26 (0.1) | 0.48 | 8.838 (18.1) | **<0.001** |
| 15 mg /1 mg | 11 | 0.25 (0.1) | 0.44 | 7.266 (10.0) | **<0.001** |
| Generic, ng/mL | 50 | 3.25 (1.8) | 0.54 | 12.213 (45.4) | **0.022** |
| 2 mg | 20 | 1.99 (0.8) | 0.42 | 10.711 (19.0) | **<0.001** |
| 4 mg | 14 | 3.24 (1.2) | 0.36 | 10.319 (13.0) | **<0.001** |
| 6 mg | 14 | 4.59 (1.7) | 0.37 | 8.144 (8.9) | **<0.001** |
| 8 mg | 2 | 6.55 (2.2) | 0.33 | - | - |
| Generic /1 mg | 50 | 0.87 (0.4) | 0.42 | 16.302 (41.7) | **<0.001** |
| 2 mg /1 mg | 20 | 1.00 (0.4) | 0.42 | 10.711 (19.0) | **<0.001** |
| 4 mg /1 mg | 14 | 0.81 (0.3) | 0.36 | 10.384 (13.0) | **<0.001** |
| 6 mg /1 mg | 14 | 0.77 (0.3) | 0.38 | 8.184 (8.9) | **<0.001** |
| 8 mg /1 mg | 2 | 0.82 (0.3) | 0.33 | - | - |

*CV: coefficient of variation; df: degrees of freedom; SD = standard deviation.*

*Note: p-values were derived from linear mixed-effects model testing the mean (intercept) against zero while accounting for repeated measures of participants.*

| **Supplementary Table S4.1. Effect of dosage on blood rapamycin levels** | | | | | | | |
| --- | --- | --- | --- | --- | --- | --- | --- |
| **Parameter** | **Estimate** | **SE** | **df** | **t** | **p-value** | **95% CI Lower** | **95% CI Upper** |
| **1 Compounded** |  |  |  |  |  |  |  |
| Intercept | -0.220 | 3.388 | 16.403 | -0.065 | 0.949 | -7.389 | 6.949 |
| **Dosage** | **0.173** | **0.071** | **25.584** | **2.442** | **0.022** | **0.027** | **0.318** |
| BMI | 0.005 | 0.106 | 18.456 | 0.045 | 0.964 | -0.217 | 0.227 |
| Sex=1 | -0.051 | 0.646 | 17.095 | -0.079 | 0.938 | -1.415 | 1.321 |
| Sex=2 | - | - | - | - | - | - | - |
| Age | 0.015 | 0.029 | 17.207 | 0.537 | 0.598 | -0.045 | 0.076 |
| **2 Generic** |  |  |  |  |  |  |  |
| Intercept | -4.220 | 1.984 | 42.269 | -2.127 | 0.039 | -8.223 | -0.217 |
| **Dosage** | **0.697** | **0.111** | **38.548** | **6.269** | **<0.001** | **0.472** | **0.922** |
| BMI | 0.109 | 0.047 | 43.846 | 2.293 | 0.027 | 0.013 | 0.204 |
| Sex=1 | 0.335 | 0.459 | 41.576 | 0.732 | 0.469 | -0.590 | 1.261 |
| Sex=2 | - | - | - | - | - | - | - |
| Age | 0.033 | 0.020 | 39.213 | 1.659 | 0.105 | -0.007 | 0.072 |
| **Supplementary Table S4.2. Interaction of dosage by formulation** | | | | | | | |
| **Parameter** | **Estimate** | **SE** | **df** | **t** | **p-value** | **95% CI Lower** | **95% CI Upper** |
| Intercept | -3.173 | 1.730 | 65.91 | -1.834 | 0.071 | -6.627 | 0.282 |
| Dosage | 0.687 | 0.111 | 65.139 | 6.169 | <0.001 | 0.464 | 0.909 |
| Group=1 | 0.401 | 0.903 | 75.224 | 0.444 | 0.658 | -1.398 | 2.201 |
| Group=2 | - | - | - | - | - | - | - |
| BMI | 0.086 | 0.043 | 72.099 | 1.983 | 0.051 | 0.000 | 0.173 |
| Sex=1 | 0.183 | 0.370 | 61.617 | 0.496 | 0.621 | -0.555 | 0.922 |
| Sex=2 | - | - | - | - | - | - | - |
| Age | 0.026 | 0.016 | 59.321 | 1.624 | 0.110 | -0.006 | 0.058 |
| **Group=1 * Dosage** | **-0.524** | **0.130** | **78.628** | **-4.026** | **<0.001** | **-0.783** | **-0.265** |
| Group=2 * Dosage | - | - | - | - | - | - | - |
| **Supplementary Table S4.3. Normalized interaction of dose and formulation** | | | | | | | |
| **Parameter** | **Estimate** | **SE** | **df** | **t** | **p-value** | **95% CI Lower** | **95% CI Upper** |
| Intercept | 0.020 | 0.414 | 58.923 | 0.048 | 0.962 | -0.808 | 0.847 |
| Dosage | -0.038 | 0.026 | 67.189 | -1.456 | 0.150 | -0.090 | 0.014 |
| **Group=1** | **-0.600** | **0.166** | **73.600** | **-3.619** | **<0.001** | **-0.930** | **-0.270** |
| Group=2 | - | - | - | - | - | - | - |
| BMI | 0.025 | 0.010 | 60.763 | 2.463 | 0.017 | 0.005 | 0.045 |
| Sex=1 | 0.086 | 0.090 | 57.595 | 0.965 | 0.339 | -0.093 | 0.266 |
| Sex=2 | - | - | - | - | - | - | - |
| Age | 0.006 | 0.004 | 55.879 | 1.521 | 0.134 | -0.002 | 0.014 |
| **Group=1 * Dosage** | **0.025** | **0.028** | **77.956** | **0.911** | **0.365** | **-0.030** | **0.081** |
| Group=2 * Dosage | - | - | - | - | - | - | - |

*CI: confidence interval; df: degrees of freedom; SE: standard error*
