## Supplementary Figure Legends for "The bioavailability of compounded and generic rapamycin in normative aging individuals: A retrospective study and review with clinical implications"

**Supplementary Figure S1. Comparative demographic distributions across rapamycin formulation types.** Real world recruitment of rapamycin users to this study resulted in **(a)** participants in the compounded rapamycin formulation group being of significantly higher age (mean = 61.7 years, SD = 9.1) than those in the generic rapamycin formulation group (mean = 56.8 years, SD = 9.3; t(65) = -2.050, p = 0.044). However, distribution of **(b)** BMI and **(c)** gender were not statistically different between formulation groups (compounded mean BMI = 23.8 kg/m2, SD: 2.9; generic BMI = 25.1 kg/m2, SD: 4.8; t(65) = 1.158, p = 0.251; compounded group males = 73.9%, generic group males = 59.1%; **^2^** (1, N = 67) = 1.443, p = 0.230). *p < 0.05 *BMI: body mass index; SD: standard deviation.*

**Supplementary Figure S2. Heterogeneity of normalized dose-to-blood level bioavailability.**

Repeated measures of the same dose in the same individuals showed consistency in bioavailability for normalized doses **(a)**, and increasing doses in the same individuals tended to increase bioavailability for normalized doses **(b)**, with no meaningful differences between formulations.
