## Supplementary figures and images for "The bioavailability of compounded and generic rapamycin in normative aging individuals: A retrospective study and review with clinical implications"

### Supplementary Figure S1

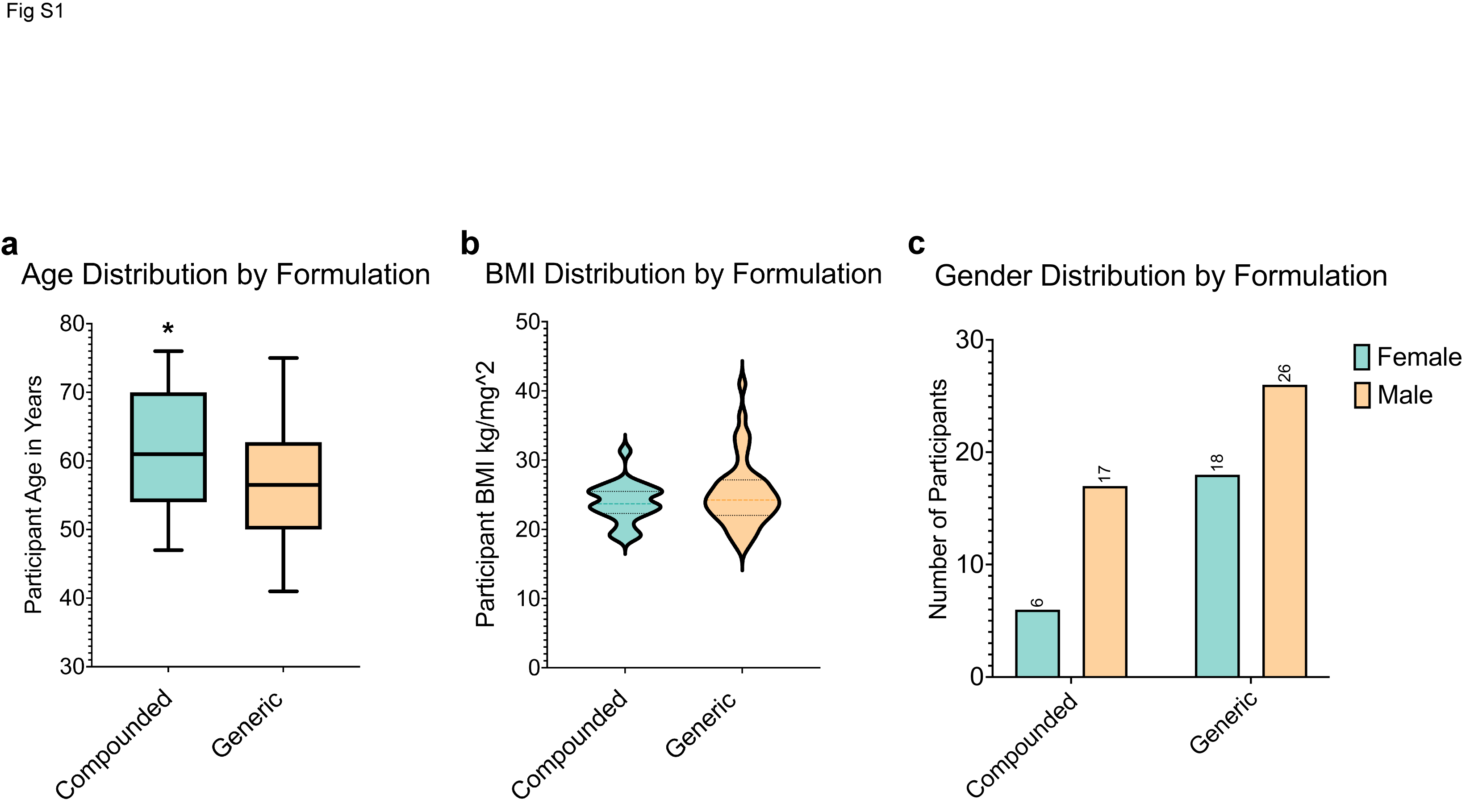

### Supplementary Figure S2

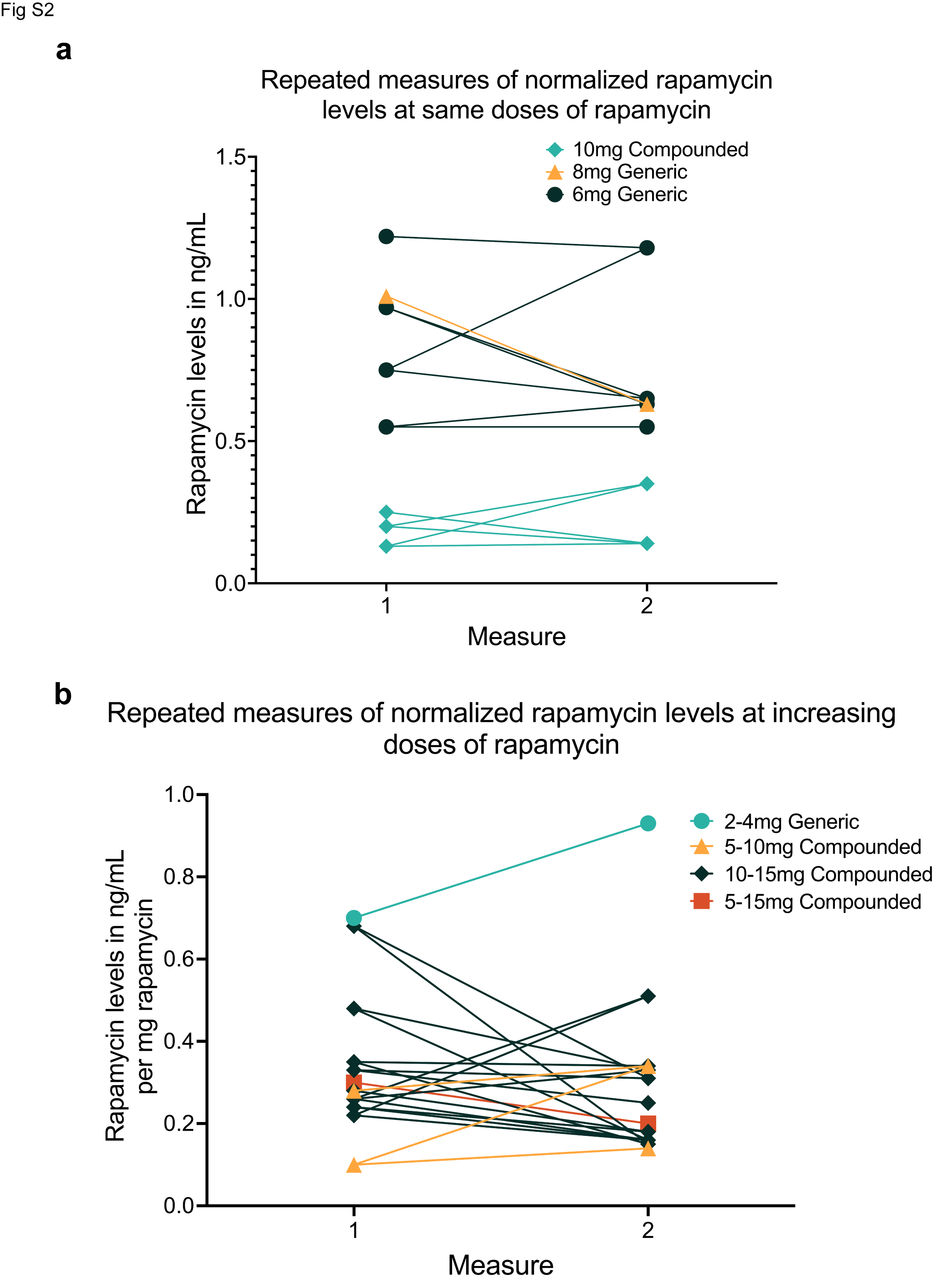
